# Towards Optimum Reporting of Pulmonary Effectiveness Databases and Outcomes (TORPEDO): identifying a core dataset for asthma and COPD studies

**DOI:** 10.1101/2021.10.14.21264843

**Authors:** Job FM van Boven, Sarah J Lucas, Gary Parker, Alan Kaplan, Antigona Trofor, Billie Bonevski, Bruce J Kirenga, Dermot Ryan, Emilio Pizzichini, Eric van Ganse, Erick Wan-Chun Huang, Evelyn Brakema, Gillian Gould, Janwillem Kocks, Jennifer Alison, Jennifer K Quint, Joan B Soriano, John Hurst, Kamran Siddiqi, Katherine Boydell, Marc Miravitlles, Mario Alberto Flores-Valdez, Marise Kasteleyn, Mark FitzGerald, Melanie Boeckmann, Michael Chaiton, Miguel Roman Rodriguez, Muralidhar Kulkarni, Nicolas Roche, Niels Chavannes, Nikolaos G Papadopoulos, Panagiotis Behrakis, Sarah Dennis, Shalini Bassi, Siân Williams, Toby M Maher, Trishul Siddharthan, Veena Kamath, Katia MC Verhamme, on behalf of REG/GACD TORPEDO collaborators

**Author notes:** **Corresponding author**: Job van Boven. University of Groningen, University Medical Center Groningen, Netherlands.

## Abstract

**Purpose:** There remains a need for a standardized dataset for respiratory studies to accelerate data collection, improve research efficiency and aid the sharing, merging and comparison of datasets. This TORPEDO (Towards Optimum Reporting of Pulmonary Effectiveness Databases and Outcomes) project aimed to develop a checklist of optimum and minimum variables for asthma and chronic obstructive pulmonary disease (COPD) research.

**Methods:** A 3-phase modified Delphi survey was conducted: in phase 1, an expert panel generated a list of variables, in phase 2 a Delphi panel selected the minimum variables (>66% agreement) for any design and in phase 3 they were asked to select a minimum set for specific study designs.

**Results:** In phase 1 the expert panel (n=22) proposed 224 variables. In phase 2, voting by 64 participants resulted in consensus (>66% agreement) for 18 variables and partial agreement (50-66%) for 44 variables, following this, 5 technical variables (e.g. date of test) were removed. In phase 3, 34 members of the Delphi panel completed voting; consensus was reached for 13 variables for retrospective asthma studies and 34 for prospective asthma studies. For COPD, there were 16 variables for retrospective studies and 37 for prospective studies. Gender, asthma/COPD exacerbations and patient-reported outcomes were the only variables with 100% agreement for both asthma and COPD studies.

**Conclusion:** The proposed list of minimally required variables will allow the assessment of current data sources for their utility in asthma and COPD studies, facilitate the merging of datasets, aid standardization of data collection and improve research efficiency.

## Introduction

Randomized controlled trials (RCTs) are considered the gold standard to assess efficacy, and, to a limited extent, the safety of a drug or non-pharmacological asthma or chronic obstructive pulmonary disease (COPD) treatment. However, because of the stringent methodology adopted with strict inclusion and exclusion criteria seen in RCTs, the relatively small sample size and short duration of follow-up, observational studies and pragmatic trials are required to provide additional information on the effectiveness and safety of a drug when used in real-life circumstances.^1-3^ Electronic health care databases, claims databases and drug and/or patient registries are important data sources for real-life studies, which has recently been underscored by regulatory authorities such as the European Medicines Agency (EMA) and the UK’s National Institute for Health and Care Excellence (NICE).^4^ With the increasing digitalization of healthcare data collection, the number of sources available for observational studies is growing exponentially, allowing research centers to conduct multinational, multi-database studies.

Heterogeneity of database structures, and differences in disease and drug coding complicates the conduct of these studies.^5^ These concerns about heterogeneity not only hold for databases, but also for the choice and definition of outcomes which makes a comparison between studies - not to mention pooling of data - difficult. The importance of choosing realistic and clinically meaningful outcomes has been described for various clinical domains including the field of respiratory research. Indeed, in 2008, the *European Respiratory Journal* published the recommendations of an (American Thoracic Society/ European Respiratory Society (ATS/ERS) task force on appropriate outcomes for COPD pharmacological trials, but no priority-analysis was performed.^6^

The Respiratory Effectiveness Group (REG, https://regresearchnetwork.org/) is an international network of respiratory experts that aims to set standards and best practices to improve real-world respiratory research. REG has made efforts to set standards for real-life respiratory research in general, but not yet at the level of specific variables.^7^ Thus, there is still an urgent need to standardize outcomes and develop a core dataset for asthma and COPD studies to accelerate data collection, improve research efficiency, replicability and transparency and create the possibility to share, compare and merge datasets around the globe to permit further analysis.^8^

The Global Alliance on Chronic Diseases (GACD) is a network of the world’s biggest public research funding agencies and funds joint programs on chronic diseases. Together, the members of the alliance represent over 90% of public health research funding worldwide. GACD aims to facilitate research collaboration on chronic disease globally with a focus on collaborations between low- and middle-income countries and vulnerable populations within high-income countries.^9^ The network coordinates and supports research activities that address the prevention and treatment of chronic non-communicable diseases, on a global scale. The work of GACD on the standardization of outcomes and development of dictionaries for chronic diseases is crucial to reach these objectives. As part of a joint effort between the REG and GACD, the TORPEDO (Towards Optimum Reporting of Pulmonary Effectiveness Databases and Outcomes) project aimed to develop a checklist with optimum and minimum required variables for research in asthma and COPD.

## Methods

### Study design

This study followed the process of a three-phased modified Delphi process^10^ as outlined in Figure 1. For this Delphi survey, various partners involved in respiratory research (clinicians, regulators, respiratory societies, patient organizations, researchers) were asked to complete a checklist asking what they considered to be optimum and minimum required variables for asthma and COPD research followed by a prioritization process of multiple rounds to reach consensus on the list of variables. The method was based on previous studies that aimed to reach consensus on a core set of items required for proper reporting of datasets.^11,12^ A point of difference to the classic Delphi approach was that, in phase 1, a smaller selected expert panel was consulted instead of the full Delphi panel to first expand the initial list with additional variables for an optimum asthma/COPD dataset. From phase 2 onwards, a full Delphi panel was established, and a prioritization process followed. The process was conducted between March 2018 and April 2019. All phases are further detailed in the next sections.

**Figure 1:**
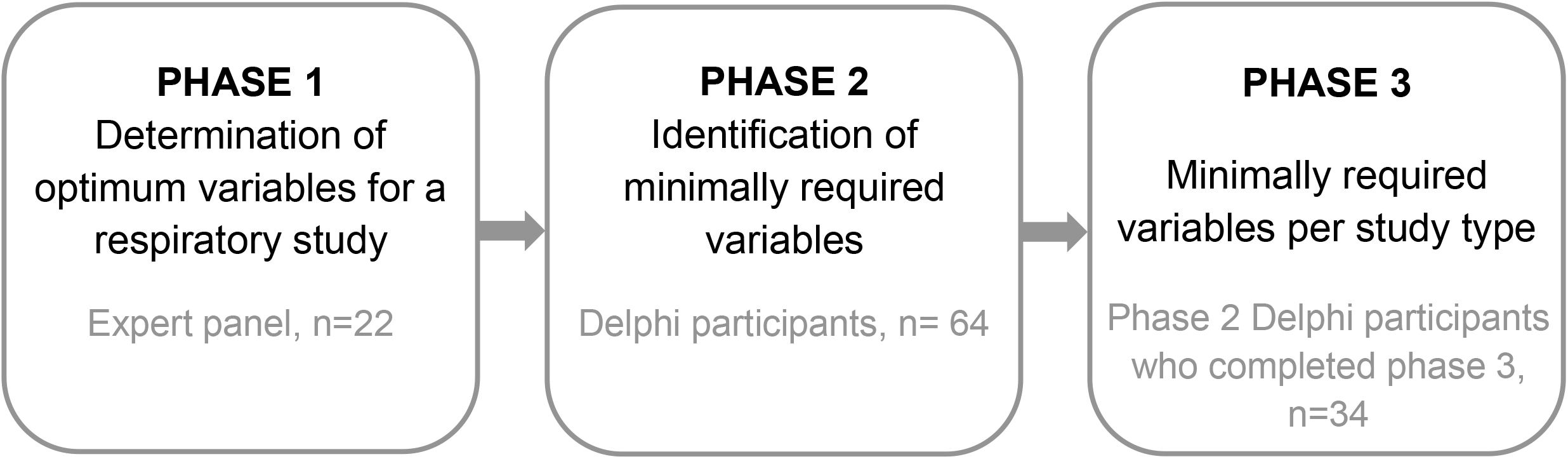
Overview of the Delphi phases

This study involved no patients; participants were all professionals involved in respiratory research who are members of the REG or GACD networks and consented to participate in the Delphi exercise. The study received ethical approval from the Anonymised Data Ethics and Protocol Transparency (ADEPT) Committee (approval number ADEPT0921).

### Outcomes

We defined a ‘minimum’ research dataset to be the minimum amount of clinical data variables to be collected to establish the effects of an intervention related to the prevention and management of asthma and COPD. An ‘optimum’ set would be the larger set of clinical data variables that could be collected where time and other related resources related to their collection and analysis were less constrained. This will provide researchers with a benchmark of which variables to select given their research goals, anticipated design and resources available.

### Phase 1

#### Identifying an optimum asthma/COPD dataset

The authors started with an initial set of variables that had been used in previous asthma/COPD studies, which were selected at the authors’ discretion. This list was circulated among a panel of respiratory experts, who could complement it with relevant variables according to their perspective. A requirement for this expert panel was that it had representation of: (i) researchers and clinicians with expertise in asthma, COPD, allergy, primary care, epidemiology and/or health economics, (ii) each continent and (iii) representatives of the major international asthma and COPD guidelines committees (Global Initiative for Asthma (GINA), Global Initiative for Chronic Obstructive Lung Disease (GOLD)). These members were all recruited through the REG (https://www.regresearchnetwork.org/). Once the list was expanded, the full list of all variables was discussed at the REG Summit (London, UK, 2017) and, if required, some variables were merged or re-categorized.

### Phase 2

In phase 2, the expert panel from phase 1 was expanded to the formal Delphi panel with more global representatives, focusing on representatives from low- and middle-income countries. These additional members were recruited through the GACD respiratory disease program. By combining the networks of REG and GACD we aimed for a representative Delphi panel where we considered the following requirements:

- High-income and low-income country representation
- Good geographical representation (all World Health Organisation regions represented, i.e. Africa, Americas, South East Asia, Europe, Eastern Mediterranean and Western Pacific)
- Wide range of researchers and clinicians with expertise in asthma, COPD, allergy, primary care, epidemiology and/or health economics
- Representation from asthma and COPD guideline committees (GINA, GOLD)

#### Voting and endorsing of variables to reduce the list to a ‘minimum’ dataset

In phase 2 of this project, all variables identified in phase 1 were presented to the Delphi panel through an online voting platform (Survey Monkey; www.surveymonkey.com). For each variable, the Delphi panel members were requested to indicate if they considered the variable an absolutely required (minimum) variable for any asthma/COPD research (no matter the design of the study, as specific design was addressed in phase 3). Variables that reached at least >66% agreement between respondents were included in the list and were moved to the next Delphi round (phase 3). Variables with partial agreement (50-66%) were further discussed at REG (Paris, France, 2018) and GACD meetings (Sao Paulo, Brazil, 2018) and if deemed relevant by the majority, included for the next Delphi round (phase 3). The 66% and 50-66% cut-off criteria were based on a previous modified Delphi exercise in severe asthma^12^.

### Phase 3

#### Prioritization of the minimum dataset per disease and study design

In this round, a prioritization of the variables identified in phase 2 was made. The Delphi panel was provided with a survey sent out through the same online voting platform. The Delphi members were presented with the list of variables identified during phase 2. Respondents were asked to indicate which variables were a minimum requirement for the study designs below:

- Prospective clinical asthma (field) study with original data collection
- Prospective clinical COPD (field) study with original data collection
- Retrospective asthma database study
- Retrospective COPD database study

As for phase 2, variables were included in the list if at least >66% agreement between respondents was reached. This exercise resulted in four final lists of minimally required variables for four types of respiratory studies.

## Results

An overview of variable selection is provided in Figure 2 and detailed in the following subsections by phase.

**Figure 2:**
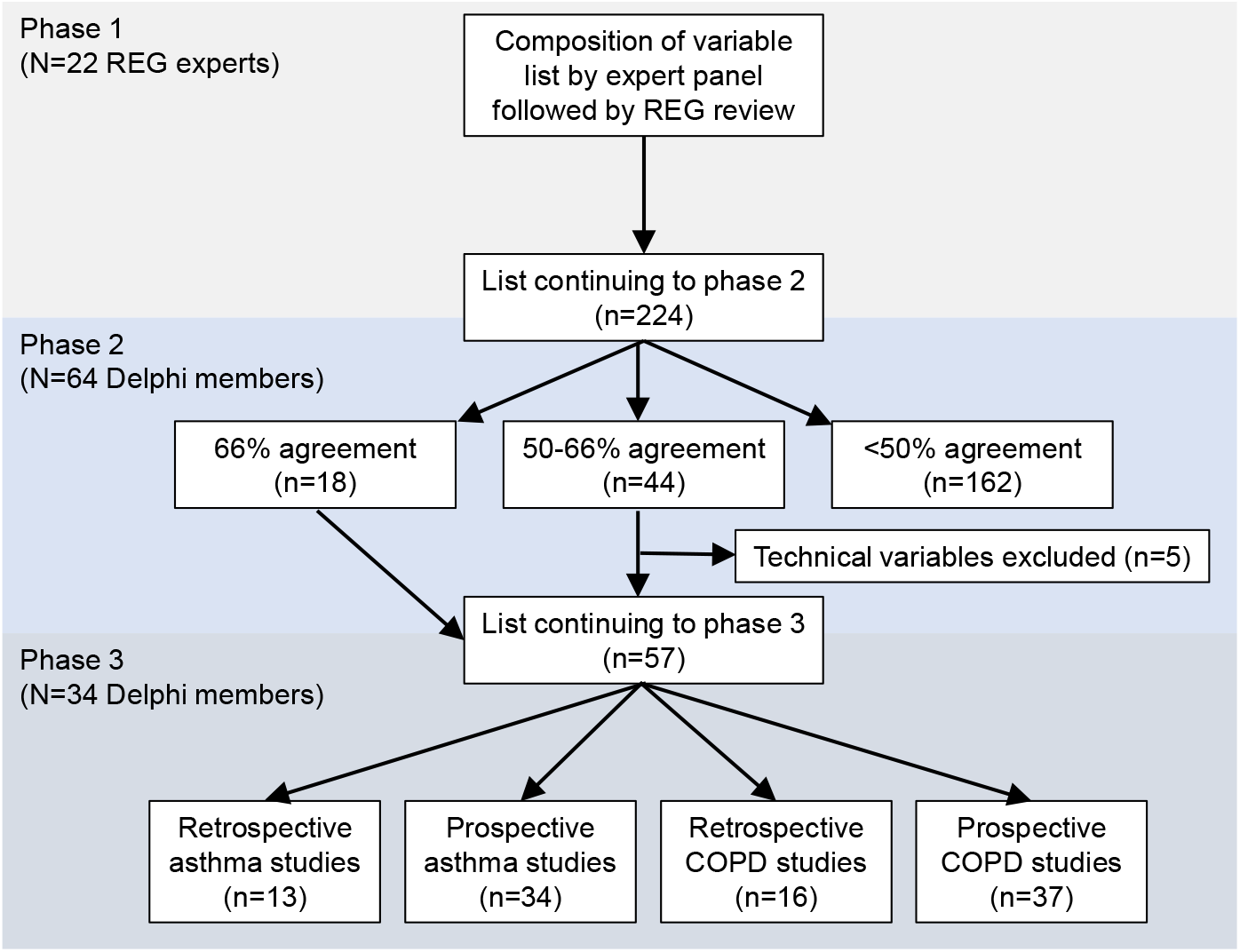
Variable flow diagram. *REG: Respiratory Effectiveness Group, COPD: Chronic Obstructive Pulmonary Disease*

### Phase 1

#### Expert panel and optimum variable list

After the suggestions of the expert panel (N=22) and REG meeting discussions, an initial list of 224 variables was generated. This list consisted of all variables deemed relevant for asthma/COPD research and is presented in Supplementary Table 1 by domain. This list was presented to the Delphi panel consulted in phase 2.

### Phase 2

#### Delphi panel

The characteristics of the Delphi panel are presented in Table 1. There was more representation from high-income than lower/middle-income countries. The majority of Delphi members had (clinical) academic positions and most indicated they had primarily in the asthma or COPD expertise, but there was also strong representation from epidemiology experts.

**Table 1:** Characteristics of the Delphi panel participants in phase 2 *(N=*64)

| **Characteristic** | **N (%)** |
| --- | --- |
| Male | 40 (63%) |
| Female | 24 (38%) |
| **Income** |  |
| High-income country | 42 (66%) |
| Low and middle income country | 22 (34%) |
| **World Health Organization region** |  |
| African region | 6 (9%) |
| Americas | 13 (20%) |
| South East Asia (inc. India, Indonesia, Nepal) | 8 (13%) |
| Europe | 26 (41%) |
| Eastern Mediterranean | 0 (0%) |
| Western Pacific (inc. Australia, China, Korea) | 11 (17%) |
| **Main workplace** |  |
| Academia | 42 (66%) |
| Hospital | 11 (17%) |
| General practice | 3 (5%) |
| Industry | 2 (3%) |
| Regulator | 1 (2%) |
| Other | 5 (8%) |
| **Core area of expertise (self-indicated)** |  |
| Asthma | 9 (14%) |
| COPD | 12 (19%) |
| Primary care | 8 (13%) |
| Pediatrics | 2 (3%) |
| Allergy | 1 (2%) |
| ILD/IPF | 2 (3%) |
| Epidemiology | 12 (19%) |
| Health economics | 6 (9%) |
| Other | 12 (19%) |

ILD: interstitial lung disease; IPF: idiopathic pulmonary fibrosis

#### List of minimum respiratory variables (uncategorized)

A total of 64 Delphi members participated in phase 2. By means of online voting on the initial set of 224 variables, immediate consensus (ie >66% agreement) was reached for 18 variables (8%). These variables are indicated by the dark grey fill in Supplementary Table 1. Partial agreement (50-66%) was reached for 44 variables (20%). These variables are indicated by the light grey fill in Supplementary Table 1. The latter were discussed at the REG (Paris) and GACD (Sao Paulo) 2018 meetings and most were deemed relevant but some only for specific studies (eg retrospective database only or prospective only) or only in specific type of studies. After phase 2, a list of 62 variables remained. After discussion, generic “technical variables” (date of test, units) were removed resulting in a set of 57 variables to be considered in phase 3.

### Phase 3

In total, 64 invitations to participate in phase 3 were sent out to the Delphi panel of which 39 members replied (61%). Of these, 5 people did not select any of variables and were hence excluded from the analysis, thus the final participants were 34 (53%).

#### Final list of minimum variables by disease and study design

After voting in this phase, from the total of 57 variables remaining from phase 2, 13 core variables were considered for retrospective asthma studies and 34 for prospective asthma studies. For COPD, this was 16 variables for retrospective studies versus 37 for prospective studies. In prospective studies, the additional variables required were mainly biomarkers, lung function measurements, and variables providing further information concerning healthcare utilization and reason for visit. Of note, gender, asthma/COPD exacerbations and a relevant patient-reported outcome (Asthma Control Questionnaire (ACQ), COPD Assessment Test (CAT)) were the only variables with 100% agreement for both asthma and COPD studies. The exact percentages of agreement per variable is provided in Supplementary Table 2. The final list of variables per disease (asthma, COPD) and study design (retrospective, prospective) is provided in Table 2 and Figure 3.

**Table 2:** Final list of minimum variables, indicated by **V**, by disease and study design

|  | **Suggested format** | **Asthma**<br>**Retrospective** | **Asthma**<br>**Prospective** | **COPD**<br>**Retrospective** | **COPD**<br>**Prospective** |
| --- | --- | --- | --- | --- | --- |
| **Demographics** |  |  |  |  |  |
| Date of birth | DD-MM-YYYY | V | V | V | V |
| Gender | M/F | V | V | V | V |
| Geographical location | Country/region |  | V |  | V |
| **Clinical** |  |  |  |  |  |
| Height | Meters |  | V |  | V |
| Weight | Kg |  |  |  | V |
| Body Mass Index | Kg/m<sup>2</sup> |  | V |  | V |
| Environmental exposures | Job type |  | V |  | V |
| Smoking status | Current/former/ never | V | V | V | V |
| Pack years | Packyears | V | V | V | V |
| **Biomarkers** |  |  |  |  |  |
| FBC with differentiation | Eosinophils, neutrophils etc. |  | V |  | V |
| Immunoglobulin E | U |  | V |  |  |
| **Lung function** |  |  |  |  |  |
| FEV1_prebronchodilation | Liters |  | V |  | V |
| FEV1_postbronchodilation | Liters |  | V |  | V |
| FEV1% predicted | % |  | V | V | V |
| FVC | Liters |  | V |  | V |
| **Pollution** |  |  |  |  |  |
| Allergen exposure | Yes or no |  | V |  |  |
| Occupational exposure | " |  |  |  | V |
| **Healthcare utilization** |  |  |  |  |  |
| Primary care consultation | Number/date of visits | V | V | V | V |
| Reason Primary Care Visit | Diagnosis |  | V |  | V |
| Secondary care visit | Date and number |  | V |  | V |
| Hospitalisation | Date and number | V | V | V | V |
| Reason hospitalisation | Diagnosis |  | V |  | V |
| ER admission | Date and number | V | V | V | V |
| Reason ER admission | Diagnosis |  | V |  | V |
| ICU stay | Date and number |  | V |  | V |
| **Medication** |  |  |  |  |  |
| Generic drug name | Name | V | V | V | V |
| Frequency of use | Date and number |  | V |  | V |
| **Comorbidities** |  |  |  |  |  |
| Diabetes Mellitus | Yes or No |  |  | V | V |
| Rhinitis | " | V | V |  |  |
| Lung cancer | " |  |  | V | V |
| Bronchiectasis | " |  |  |  | V |
| Asthma | " |  |  | V | V |
| COPD | " | V | V |  |  |
| Cardiovascular disease | " |  |  | V | V |
| **Mortality** |  |  |  |  |  |
| Mortality | Life status (dead/alive) | V | V | V | V |
| Cause of death | Diagnosis |  | V |  | V |
| Primary Cause of death | Diagnosis |  | V |  | V |
| Date of death | Date | V | V | V | V |
| **Exacerbations** |  |  |  |  |  |
| Asthma exacerbations | Date and number | V | V |  |  |
| COPD exacerbations | " |  |  | V | V |
| Other severe respiratory episodes | " |  | V |  | V |
| **Patient reported outcomes** |  |  |  |  |  |
| ACQ | Score |  | V |  |  |
| CAT | " |  |  |  | V |

ACQ: asthma control questionnaire; CAT: COPD Assessment Test; ER: emergency room; FBC: full blood count; FEV1: forced expiratory volume in 1 second; FVC: forced vital capacity; ICU: intensive care unit.

**Figure 3:**
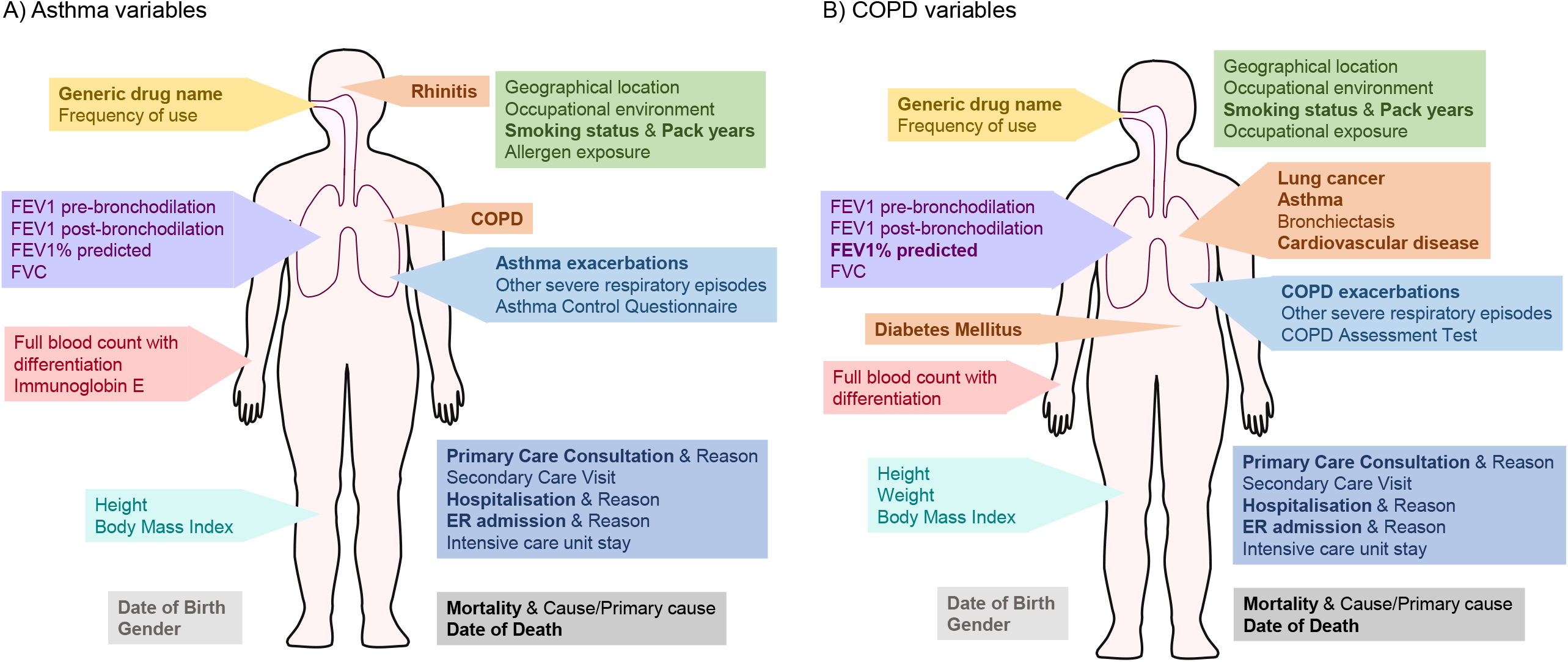
Overview of REG/GACD core dataset for prospective asthma (A) and COPD (B) studies, with variables for retrospective studies indicated in bold. *COPD: chronic obstructive pulmonary disease; ER: emergency room; FEV1: forced expiratory volume in 1 second; FVC: forced vital capacity*.

## Discussion

We used a modified Delphi exercise to determine a recommended core dataset of minimum variables for asthma and COPD studies. Following the identification of the full list of potential asthma/COPD variables by a panel of 22 REG experts, 64 participants completed phase 2 to determine the minimally recommended variables, 34 of these completed phase 3 to determine the minimal variables per study type. There was generally good agreement concerning the variables needed for retrospective studies, regardless of whether the study was investigating asthma or COPD; similarly, for prospective studies, the variables required were very similar for both COPD and asthma studies. For prospective studies, where new data collection would be possible, the participants felt that a greater number of variables should be required than for retrospective studies. In prospective studies, the additional variables required were mainly biomarkers, lung function measurements, and variables giving further information concerning healthcare utilization and reason for visit. The fewer required variables in retrospective studies likely reflects the experience of the participants who were aware of the limitations of retrospective data sources. Indeed, the accuracy and regularity of recording of variables might influence a participant’s decision on whether that variable should be included, and even its utility.

To facilitate database studies, but also for existing longitudinal cohorts, there is a need to determine whether databases contain the minimum required variables to adequately conduct the study. Often in observational database studies, particularly for rare events and conditions, it is beneficial to include data from multiple data sources to increase the power. Also, the ability to pool data from multiple datasets allows a more heterogeneous population of patients to be included from different cultural backgrounds. The Uncovering and Noting Long-term Outcomes in COPD and asthma to enhance Knowledge (UNLOCK) initiative, created in 2010, aimed to determine the minimum required dataset for observational studies in asthma and COPD management in primary care, which is an important step to enable pooling of data from multiple datasets.^13^ Indeed, the UNLOCK group subsequently found that a key challenge in using real-world data from across different countries and regions was the lack of comparability between datasets, with not all of them containing all the required data variables or having the same definitions of the same variable.^14^ A difference with our study is that in our study a structured Delphi approach was used (compared to a focus group discussion in the UNLOCK study) and that we expanded our research beyond primary care and existing datasets.

Data reuse can be severely hampered when datasets are not standardized. Therefore, it is important to ensure the minimum required variables are included in new (thus prospective) data collection.^15^ The Core Outcome Measures in Effectiveness Trials (COMET) Initiative also aims to develop standardized sets of outcomes, and these sets should be considered the minimum variables that should be measured and reported in clinical trials.^15^ Importantly, such lists did not yet exist for asthma and/or COPD and needed to be developed for use beyond clinical trials only. The development of core datasets is also important to understand the implementation of interventions. Where we have standardized datasets, we are more likely be able to understand differences between centers as a result of true differences rather than differences in variables.

Several groups have previously considered defining core variables or have developed a list of minimum required variables using a Delphi approach. A similar style of Delphi exercise was used to determine the minimum variables required when creating the International Severe Asthma Registry (ISAR), which reached a consensus on 95 variables including patient demographics, medical history, patient-reported outcomes, diagnostic information, clinical characteristics and physician-reported outcomes.^12^ While the ATS/ERS Task Force on “Outcomes for COPD pharmacological trials: from lung function to biomarkers” did not determine a list of minimum variables they did highlight certain variables that COPD trials should include, such as lung function variables other than forced expiratory volume in 1 second (FEV1) (e.g. forced vital capacity (FVC), inspiratory capacity to total lung capacity ratio and measures of dyspnoea) and the frequency of exacerbations.^6^ Information on COPD exacerbations was included in both our retrospective and prospective COPD lists, and some lung function measures other than FEV1 were included on the prospective COPD study list. An UNLOCK study, combining data from primary care across Europe to determine the prevalence of comorbidities in COPD patients, and their impact on health status and COPD, required datasets to contain the following variables: age, gender, FEV1 and ideally FEV1/FVC, CAT or Clinical COPD Questionnaire score, Medical Research Council (MRC) dyspnea score, body mass index, smoking status, education level and comorbidities (Heart disease, hypertension, diabetes, depression and asthma).^16^ These required variables are comparable to those included in our list, but there are some differences (in comorbidities and requirement for MRC score), which are mainly due to the specific requirements of the UNLOCK study. Indeed, the list produced here provides purely the core recommended variables; in practice the minimum variables required for a particular study will depend on the specific question to be addressed.

Pooling of data, not only from clinical trials but also with regard to real-world observational studies becomes increasingly more important, and harmonization of data by means of a common data model not only optimizes data extraction but also data pooling.^17^ Ideally, the dictionary used by the common data model should also include the minimum variables as suggested in our retrospective asthma and COPD lists.

One of the strengths of this study is that the Delphi panel (n=64) comprised of a wide range of participants from: high- and low and-middle income countries, different geographic regions, different areas of expertise and different professional backgrounds. However, only 34 participants (53%) participated in both phase 2 and 3, and the reason to abstain for phase 3 is not known. This is, however, a common challenge in online Delphi studies and given we have no reason to assume that biased exclusion occurred (“missing at random”), we feel that it did not significantly impact the validity of our results.

While it has been determined that some variables such as the full date of birth were required, often local regulatory/legal rules such as General Data Protection Regulation (GDPR) might not allow inclusion of potential identifying patient information.^18^ Therefore, other formats or related variables may need to be used, such as only including year of birth or age at the date of enrolment.

This list is valid at the present time but may require updating in the future as the data required changes over time with increased knowledge giving a better understanding of risk factors, improved diagnostic criteria and clinical management. For example, FEV1 measurements are currently key in diagnosing COPD, however, it is hoped in the future there may be validated biomarkers for COPD, which may need to be included in a future list of minimum required dataset.^6^

Lastly, there remains debate around the precise definition of some disease terms, for example exacerbations, and for standardization across studies it is important that the same definition has been used.

Determined by a global Delphi panel of 64 participants, these proposed minimum required variables will facilitate the assessment of current data sources for their potential utility for use in asthma and COPD studies. It provides a basis to aid the standardization of data collection and improve research efficiency, replicability and transferability. Determining these minimal variables is an important step in facilitating the sharing, comparing and merging of datasets.

## Data Availability

All data produced in the present work are contained in the manuscript

