## Supplementary tables for "Towards Optimum Reporting of Pulmonary Effectiveness Databases and Outcomes (TORPEDO): identifying a core dataset for asthma and COPD studies"

**Supplementary table 1**. List of optimum variables for respiratory research, determined by the expert panel and presented to the Delphi participants in phase 2. The number and percentage of participant who considered the variable to be absolutely required/minimum variable for any asthma/COPD research are given. Dark grey fill indicates variables with >66% agreement, and lighter grey fill indicates variables with 50-66% agreement.

| Survey Variable | No. of participants, n = 64 | % of participants |
|---|---|---|
| **Demographics** | | |
| Date of birth | 54 | 84.4 |
| Time of birth | 1 | 1.6 |
| Details of birth | 17 | 26.6 |
| Gender | 57 | 89.1 |
| Nationality | 25 | 39.1 |
| Ethnicity | 44 | 68.8 |
| Marital status | 20 | 31.3 |
| Number of dependents | 10 | 15.6 |
| Number of people in household | 26 | 40.6 |
| Level of education | 37 | 57.8 |
| Socio-economic status | 45 | 70.3 |
| Geographical location | 32 | 50.0 |
| Salary range | 13 | 20.3 |
| Health insurance coverage | 25 | 39.1 |
| Place of birth | 13 | 20.3 |
| Occupation/Working environment | 46 | 71.9 |
| Family/Household income | 29 | 45.3 |
| Height | 44 | 68.8 |
| Weight | 46 | 71.9 |
| Obesity | 8 | 12.5 |
| Body Mass Index | 34 | 53.1 |
| Alcohol use | 35 | 54.7 |
| Illicit drug use | 28 | 43.8 |
| Diet | 23 | 35.9 |
| Smoking - current status | 53 | 82.8 |
| Smoking - pack years | 41 | 64.1 |
| Smoking - no. of cigarettes | 28 | 43.8 |
| Smoking - years exposed | 28 | 43.8 |
| Smoking - parental smoking | 30 | 46.9 |
| Smoking - passive smoking | 35 | 54.7 |
| Physical activity | 40 | 62.5 |
| **Blood testing** | | |
| Date of test | 37 | 57.8 |
| Units | 35 | 54.7 |
| Normal range | 28 | 43.8 |
| Full blood count with differentiation | 40 | 62.5 |
| Hemoglobin | 26 | 40.6 |
| Creatinine | 22 | 34.4 |

| | | |
|---|---|---|
| Glomerular filtration rate | 12 | 18.8 |
| Liver function | 15 | 23.4 |
| C-reactive protein | 24 | 37.5 |
| Immunoglobulin E (total, specific) | 38 | 59.4 |
| Immunoglobulin G (total, specific) | 12 | 18.8 |
| Skin prick test | 13 | 20.3 |
| Dipeptidyl peptidose IV (DPPIV) | 3 | 4.7 |
| Serum periostin | 4 | 6.3 |
| Vitamin D | 11 | 17.2 |
| O2 saturation | 24 | 37.5 |
| Blood gases | 16 | 25.0 |
| Genetics/Genomic profiling | 6 | 9.4 |
| Fibrinogen, BNP, Troponin | 9 | 14.1 |
| Alpha-1-anti-trypsin | 12 | 18.8 |
| Serum MMP-7, KL6, SP-A, collagen degradation products | 4 | 6.3 |
| **Lung markers** | | |
| Date of lung function measurement | 45 | 70.3 |
| Forced Expiratory Volume in 1 second (FEV1) | 42 | 65.6 |
| Pre-broncodilator Forced Expiratory Volume in 1 second (FEV1-pre) | 35 | 54.7 |
| Post-broncodilator Forced Expiratory Volume in 1 second (FEV1-post) | 37 | 57.8 |
| Forced Expiratory Volume in 1 second % predicted (FEV1% pred) | 32 | 50.0 |
| Forced Vital Capacity (FVC) | 41 | 64.1 |
| Forced Vital Capacity % predicted (FVC%pred) | 27 | 42.2 |
| Peak Expiratory Flow (PEF) | 21 | 32.8 |
| Fraction of exhaled nitric oxide (FeNO) | 21 | 32.8 |
| Exhaled Breath Condensate | 6 | 9.4 |
| Bronchodilator response | 19 | 29.7 |
| Impulse oscillometry (IOS) | 6 | 9.4 |
| Bronchial Hyperreactivity (BHR) | 10 | 15.6 |
| Carbon monoxide (CO) | 17 | 26.6 |
| Diffusing capacity of the lungs for carbon monoxide (DLCO) | 19 | 29.7 |
| Carbon monoxide transfer coefficient (KCO) | 12 | 18.8 |
| Total Lung Capacity (TLC) | 16 | 25.0 |
| Broncho-Alveolar Lavage (BAL) | 6 | 9.4 |
| Poligraphy | 5 | 7.8 |
| Audio Assessment (e.g. assessment for lung crackles/Velcro) | 5 | 7.8 |
| Bronchoscopy | 6 | 9.4 |
| Bronchoscopy transbronchial lung biopsy | 6 | 9.4 |
| Exercise test (e.g. 6 minutes walk test) | 19 | 29.7 |
| Open Lung Biopsy (OLBx) [for Interstitial Lung Disease (ILD) patients] | 8 | 12.5 |
| Cough | 28 | 43.8 |
| Sputum | 25 | 39.1 |
| **Imaging** | | |
| Date of imaging | 37 | 57.8 |
| Chest X-ray | 32 | 50.0 |
| High resolution computer tomography (HRCT) | 20 | 31.3 |
| MRI | 7 | 10.9 |
| PET | 6 | 9.4 |

| | | |
|---|---|---|
| V/q scan | 6 | 9.4 |
| CT angiogram of the chest | 6 | 9.4 |
| Echocardiogram | 13 | 20.3 |
| CT scan of the thorax | 13 | 20.3 |
| **Environmental Factors** | | |
| Outdoor air pollution | 36 | 56.3 |
| Indoor/household air pollution | 40 | 62.5 |
| Allergen exposures (mould, pets) | 37 | 57.8 |
| Occupational exposure | 45 | 70.3 |
| Secondhand smoke exposure | 38 | 59.4 |
| **Healthcare Utilization** | | |
| Date of contact/contact (healthcare) | 43 | 67.2 |
| Primary Care Consultations | 44 | 68.8 |
| Reason for Primary Care Consultations | 35 | 54.7 |
| type of contacts (telephone, face-to-face, home visits …) | 15 | 23.4 |
| Secondary Care Consultations | 42 | 65.6 |
| Reason for Secondary Care Consultations (e.g. for lower respiratory complaints) | 32 | 50.0 |
| Unscheduled healthcare contacts | 31 | 48.4 |
| Hospitalisations/ Hospital admission | 47 | 73.4 |
| Hospital discharge date | 18 | 28.1 |
| Duration of hospitalisation | 29 | 45.3 |
| Reason for Hospitalisations/Hospital admission (e.e. for lower respiratory complaints) | 42 | 65.6 |
| A&E/ER attendances | 39 | 60.9 |
| Reason for A&E/ER attendances (e.g for lower respiratory complaint) | 36 | 56.3 |
| ICU Stay | 37 | 57.8 |
| Reason for ICU Stay (e.g pneumonia) | 29 | 45.3 |
| ICU duration | 23 | 35.9 |
| Out patient department attendance | 24 | 37.5 |
| Reason for outpatient department attendance (e.g. rehabilitation) | 20 | 31.3 |
| Rehabilitation | 27 | 42.2 |
| Duration of Rehabilitation | 22 | 34.4 |
| Physiotherapy | 26 | 40.6 |
| Reason for physiotherapy | 17 | 26.6 |
| Duration physiotherapy | 16 | 25.0 |
| Out of hours attendances | 13 | 20.3 |
| Personnel time | 8 | 12.5 |
| Cost of HRU | 15 | 23.4 |
| **Treatments** | | |
| Generic drug name | 39 | 60.9 |
| Brand drug name | 17 | 26.6 |
| Prescribed Drug Name | 24 | 37.5 |
| Prescribed Drug Class | 16 | 25.0 |
| Date prescribed | 33 | 51.6 |
| Dispensed Drug Name | 22 | 34.4 |
| Dispensed Drug Class | 13 | 20.3 |
| Date Dispensed | 24 | 37.5 |

| | | |
|---|---|---|
| Indication of use | 23 | 35.9 |
| Drug strength | 29 | 45.3 |
| Pack size | 20 | 31.3 |
| Frequency of Use/Prescribing instructions | 36 | 56.3 |
| Quantity of refill | 20 | 31.3 |
| Device (e.g. metered dose inhaler) | 28 | 43.8 |
| Spacer (e.g. aerochamber) | 23 | 35.9 |
| Inhaler technique | 26 | 40.6 |
| Over the counter medications | 18 | 28.1 |
| Self-management (or action) plan | 28 | 43.8 |
| Oxygen Usage (intermittent or chronic) | 26 | 40.6 |
| Peak flow metre (registration as part of patient management) | 13 | 20.3 |
| Adherence | 29 | 45.3 |
| CPAP | 17 | 26.6 |
| Non-invasive ventilation | 17 | 26.6 |
| Pneumococcal vaccination history | 28 | 43.8 |
| Influenza vaccination history | 29 | 45.3 |
| **Coding system** | | |
| ATC (Anatomical Therapeutic Chemical) | 22 | 34.4 |
| Multilex | 7 | 10.9 |
| NDC (National Drug Code - often used in the US) | 11 | 17.2 |
| J-codes for office administered drugs (e.g. biologics) | 7 | 10.9 |
| RxNorm | 10 | 15.6 |
| ICD-9 | 6 | 9.4 |
| ICD-10 | 26 | 40.6 |
| ICD-11 | 15 | 23.4 |
| ICPC | 6 | 9.4 |
| Read | 5 | 7.8 |
| SNOMED CT | 12 | 18.8 |
| **Comorbidities** | | |
| Parental Atopy | 17 | 26.6 |
| Premature birth | 18 | 28.1 |
| Charlson Comorbidity Index | 25 | 39.1 |
| Date of disease diagnosis | 25 | 39.1 |
| Reported date | 10 | 15.6 |
| Resolved date | 7 | 10.9 |
| Status (active, resolved, other) | 12 | 18.8 |
| Diabetes Mellitus | 34 | 53.1 |
| Eczema | 22 | 34.4 |
| Rhinitis | 33 | 51.6 |
| Nasal Polyps | 30 | 46.9 |
| Osteoporosis | 22 | 34.4 |
| Psychiatric disorders | 23 | 35.9 |
| Depression/Anxiety | 30 | 46.9 |
| Gastroesophageal Reflux Disease | 24 | 37.5 |
| Chronic Kidney Disease | 17 | 26.6 |
| Lung Cancer | 33 | 51.6 |
| Sleep Apnoea | 29 | 45.3 |

| | | |
|---|---|---|
| Anaemia | 18 | 28.1 |
| Cognitive Dysfunction | 19 | 29.7 |
| Cerebrovascular Disease | 23 | 35.9 |
| Bronchiectasis | 32 | 50.0 |
| Asthma | 43 | 67.2 |
| COPD | 43 | 67.2 |
| Asthma-COPD Overlap | 31 | 48.4 |
| Interstitial Lung Disease | 27 | 42.2 |
| Tuberculosis | 37 | 57.8 |
| HIV/AIDS | 30 | 46.9 |
| Cardiovascular Disease | 36 | 56.3 |
| Ischaemic Heart Disease | 30 | 46.9 |
| Hypertension | 29 | 45.3 |
| Heart Failure | 31 | 48.4 |
| Myocardial Infarction | 26 | 40.6 |
| Rheumatoid Arthritis | 24 | 37.5 |
| Nasal hyper-reactivity | 11 | 17.2 |
| Bronchial hyper-reactivity | 12 | 18.8 |
| Connective tissue disease and features suggestive of CTD (Raynaud's phenomenon, Morning stiffness, digital oedema | 15 | 23.4 |
| Sinusitis | 23 | 35.9 |
| Rhinosinusitis | 26 | 40.6 |
| Pneumonia | 31 | 48.4 |
| Osteoporosis | 18 | 28.1 |
| Other cancers besides lung | 18 | 28.1 |
| **Mortality** | | |
| Mortality Status – Alive/Deceased | 44 | 68.8 |
| Cause of death | 41 | 64.1 |
| Primary cause of death | 34 | 53.1 |
| Secondary/contributing cause of death | 27 | 42.2 |
| Date of Death | 42 | 65.6 |
| **Exacerbations** | | |
| Asthma Exacerbations | 44 | 68.8 |
| COPD Exacerbations | 43 | 67.2 |
| Interstitial lung disease exacerbations | 29 | 45.3 |
| Include other severe respiratory episodes (e.g. influenza, pneumonia) | 33 | 51.6 |
| Severity of exacerbations | 31 | 48.4 |
| CF exacerbations | 21 | 32.8 |
| **Patient Reported Outcomes** | | |
| Asthma Symptom Utility Index (ASUI) | 9 | 14.1 |
| Asthma control questionnaire (ACQ) | 32 | 50.0 |
| (Children's) asthma control test (ACT) | 25 | 39.1 |
| Asthma bother profile (ABP) | 4 | 6.3 |
| (Pediatric) asthma related quality of life questionnaire (AQLQ) | 11 | 17.2 |
| Asthma impact survey (AIS-6) | 7 | 10.9 |
| COPD assessment test (CAT) | 33 | 51.6 |
| Clinical COPD Questionnaire (CCQ) | 22 | 34.4 |
| Saint Georges Respiratory Questionnaire (SGRQ) | 21 | 32.8 |

| | | |
|---|---|---|
| Dyspnoea – Royal College of Physicians 3 Questions (RCP3) | 11 | 17.2 |
| EQ-5D | 26 | 40.6 |
| Sino-nasal outcome test (SNOT-22) | 8 | 12.5 |
| Rhinitis Control Assessment Test | 6 | 9.4 |
| Control of allergic rhinitis and asthma test (CARAT) | 9 | 14.1 |
| Hospital Anxiety depression (HADS) | 16 | 25.0 |
| (modified) Medical Research Council Dyspnea scale (MRC) | 20 | 31.3 |
| Short form health survey (SF-12) | 13 | 20.3 |

**Supplementary table 2**. Number (and percentage) of participants in phase 3 who considered each variable as required for each type of study. Grey fill indicate variables with >66% agreement between participants.

| | Asthma Retrospective | Asthma Prospective | COPD Retrospective | COPD Prospective |
|---|---|---|---|---|
| **Demographics** | | | | |
| Date of birth | **32 (94.1)** | **33 (97.1)** | **32 (94.1)** | **33 (97.1)** |
| Gender | **34 (100)** | **34 (100)** | **34 (100)** | **34 (100)** |
| Ethnicity | 11 (32.4) | 18 (52.9) | 11 (32.4) | 18 (52.9) |
| Level of education | 9 (26.5) | 16 (47.1) | 9 (26.5) | 16 (47.1) |
| Socio economic status | 13 (38.2) | 21 (61.8) | 12 (35.3) | 22 (64.7) |
| Geographical location | 18 (52.9) | **23 (67.6)** | 18 (52.9) | **23 (67.6)** |
| **Clinical** | | | | |
| Height | 19 (55.9) | **28 (82.4)** | 18 (52.9) | **28 (82.4)** |
| Weight | 14 (41.2) | 14 (41.2) | 14 (35.9) | **24 (70.6)** |
| Body Mass Index | 21 (61.8) | **24 (70.6)** | 20 (58.8) | **23 (67.6)** |
| Occupational environment | 17 (50) | **28 (82.4)** | 18 (52.9) | **28 (82.4)** |
| Alcohol use | 4 (11.8) | 8 (23.5) | 4 (11.8) | 8 (23.5) |
| Smoking status | **29 (85.3)** | **32 (94.1)** | **29 (85.3)** | **32 (94.1)** |
| Pack years | **24 (70.6)** | **30 (88.2)** | **25 (73.5)** | **32 (94.1)** |
| Passive smoking | 13 (38.2) | 22 (64.7) | 13 (38.2) | 23 (67.6) |
| Physical activity | 6 (17.6) | 19 (55.9) | 8 (23.5) | 20 (58.8) |
| **Biomarkers** | | | | |
| Full blood count with differentiation | 18 (52.9) | **26 (76.5)** | 17 (50) | **27 (79.4)** |
| Immunoglobulin E | 10 (29.4) | **27 (79.4)** | 3 (8.8) | 9 (26.5) |
| **Lung function** | | | | |
| FEV1 prebronchodilation | 15 (44.1) | **25 (73.5)** | 17 (50) | **24 (70.6)** |
| FEV1 postbronchodilation | 16 (47.1) | **28 (82.4)** | 20 (58.8) | **31 (91.2)** |
| FEV1% predicted | 21 (61.8) | **26 (76.5)** | **23 (67.6)** | **28 (82.4)** |
| FVC | 19 (55.9) | **25 (73.5)** | 21 (61.8) | **28 (82.4)** |
| **Environmental factors** | | | | |
| Outdoor air pollution | 6 (17.6) | 12 (35.3) | 8 (23.5) | 13 (38.2) |
| Indoor air pollution | 8 (23.5) | 15 (44.1) | 9 (26.5) | 15 (44.1) |
| Allergen exposure | 12 (35.3) | **23 (67.6)** | 2 (5.9) | 6 (17.6) |
| Occupational exposure | 15 (44.1) | 21 (61.8) | 17 (50) | **23 (67.6)** |
| Secondhand smoke | 11 (32.4) | 19 (55.9) | 12 (35.3) | 19 (55.9) |
| **Healthcare utilization** | | | | |
| Primary care consultation | **25 (73.5)** | **27 (79.4)** | **25 (73.5)** | **27 (79.4)** |
| Reason Primary Care Visit | 14 (41.2) | **23 (67.6)** | 14 (41.2) | **23 (67.6)** |
| Secondary care visit | 20 (58.8) | **25 (73.5)** | 21 (61.8) | **24 (70.6)** |
| Reason Secondary Care visit | 11 (32.4) | 19 (55.9) | 11 (32.4) | 19 (55.9) |

| | | | | |
|---|---|---|---|---|
| Hospitalisation | **30 (88.2)** | **31 (91.2)** | **30 (88.2)** | **31 (91.2)** |
| Reason hospitalisation | 20 (58.8) | **26 (76.5)** | 20 (58.8) | **26 (76.5)** |
| ER admission | **26 (76.5)** | **30 (88.2)** | **25 (73.5)** | **29 (85.3)** |
| Reason ER admission | 15 (44.1) | **26 (76.5)** | 14 (41.2) | **25 (73.5)** |
| ICU stay | 18 (52.9) | **23 (67.6)** | 17 (50) | **23 (67.6)** |
| Chest X-ray | 3 (8.8) | 12 (35.3) | 5 (14.7) | 17 (50) |
| **Medication** | | | | |
| Generic drug name | **32 (94.1)** | **34 (100)** | **32 (94.1)** | **34 (100)** |
| Date prescribed | 20 (58.8) | 22 (64.7) | 20 (58.8) | 22 (64.7) |
| Frequency of use | 22 (64.7) | **26 (76.5)** | 22 (64.7) | **26 (76.5)** |
| **Comorbidities** | | | | |
| Diabetes Mellitus | 18 (52.9) | 18 (52.9) | **24 (70.6)** | **27 (79.4)** |
| Rhinitis | **27 (79.4)** | **32 (94.1)** | 13 (38.2) | 17 (50) |
| Lung cancer | 14 (41.2) | 15 (44.1) | **27 (79.4)** | **32 (94.1)** |
| Bronchiectasis | 16 (47.1) | 16 (47.1) | 22 (64.7) | **26 (76.5)** |
| Asthma | 21 (61.8) | 21 (61.8) | **32 (94.1)** | **34 (100)** |
| COPD | **30 (88.2)** | **31 (91.2)** | 22 (64.7) | 22 (64.7) |
| TBC | 14 (41.2) | 14 (41.2) | 19 (55.9) | 22 (64.7) |
| Cardiovascular disease | 15 (44.1) | 16 (47.1) | **27 (79.4)** | **30 (88.2)** |
| **Mortality** | | | | |
| Mortality | **31 (91.2)** | **30 (88.2)** | **33 (97.1)** | **32 (94.1)** |
| Cause of death | 18 (52.9) | **23 (67.6)** | 19 (55.9) | **24 (70.6)** |
| Primary Cause of death | 19 (55.9) | **28 (82.4)** | 20 (58.8) | **29 (85.3)** |
| Date of death | **23 (67.6)** | **26 (76.5)** | **24 (70.6)** | **27 (79.4)** |
| **Exacerbations** | | | | |
| Asthma exacerbations | **33 (97.1)** | **34 (100)** | 12 (35.3) | 12 (35.3) |
| COPD exacerbations | 12 (35.3) | 11 (32.4) | **33 (97.1)** | **34 (100)** |
| Other severe respiratory episodes | 21 (61.8) | **25 (73.5)** | 21 (61.8) | **27 (79.4)** |
| **Patient Reported Outcomes** | | | | |
| ACQ | 19 (55.9) | **34 (100)** | 2 (5.9) | 4 (11.8) |
| CAT | 2 (5.9) | 5 (14.7) | 19 (55.9) | **34 (100)** |

ACQ: asthma control questionnaire; CAT: COPD Assessment Test; ER: emergency room; FEV1: forced expiratory volume in 1 second; FVC: forced vital capacity; ICU: intensive care unit.
